# A 2-Gene Host Signature for Improved Accuracy of COVID-19 Diagnosis Agnostic to Viral Variants

**DOI:** 10.1101/2022.01.06.21268498

**Authors:** Jack Albright, Eran Mick, Estella Sanchez-Guerrero, Jack Kamm, Anthea Mitchell, Angela M. Detweiler, Norma Neff, Alexandra Tsitsiklis, Paula Hayakawa Serpa, Kalani Ratnasiri, Diane Havlir, Amy Kistler, Joseph L. DeRisi, Angela Oliveira Pisco, Charles R. Langelier

**Affiliations:** Chan Zuckerberg Biohub, San Francisco, CA, USA; Division of Infectious Diseases, Department of Medicine, University of California San Francisco, San Francisco, CA, USA; Division of Pulmonary and Critical Care Medicine, Department of Medicine, University of California San Francisco, San Francisco, CA, USA; Department of Biochemistry and Biophysics, University of California San Francisco, San Francisco, CA, USA; Division of HIV, Infectious Diseases and Global Medicine, Department of Medicine, University of California San Francisco, San Francisco, CA, USA

## Abstract

The continued emergence of SARS-CoV-2 variants is one of several factors that may cause false negative viral PCR test results. Such tests are also susceptible to false positive results due to trace contamination from high viral titer samples. Host immune response markers provide an orthogonal indication of infection that can mitigate these concerns when combined with direct viral detection. Here, we leverage nasopharyngeal swab RNA-seq data from patients with COVID-19, other viral acute respiratory illnesses and non-viral conditions (n=318) to develop support vector machine classifiers that rely on a parsimonious 2-gene host signature to predict COVID-19. Optimal classifiers achieve an area under the receiver operating characteristic curve (AUC) greater than 0.9 when evaluated on an independent RNA-seq cohort (n=553). We show that a classifier relying on a single interferon-stimulated gene, such as *IFI6* or *IFI44*, measured in RT-qPCR assays (n=144) achieves AUC values as high as 0.88. Addition of a second gene, such as *GBP5*, significantly improves the specificity compared to other respiratory viruses. The performance of a clinically practical 2-gene RT-qPCR classifier is robust across common SARS-CoV-2 variants, including Omicron, and is unaffected by cross-contamination, demonstrating its utility for improving accuracy of COVID-19 diagnostics.

## Introduction

The COVID-19 pandemic has inflicted unprecedented human health consequences, with millions of deaths reported worldwide since December 2019^1^. Testing is a cornerstone of pandemic management, yet existing assays suffer from accuracy limitations. Even the gold-standard testing modality of nasopharyngeal (NP) swab RT-PCR returns falsely negative in a substantial proportion of cases^2–4^ and may fail to detect SARS-CoV-2 variants with mutations at primer target sites^5–7^. False positive tests due to sample cross-contamination in the laboratory are also a significant complication^8,9^ as they can lead to costly contact tracing efforts and the unnecessary isolation of uninfected individuals, including essential workers.

Measuring the host immune response offers a complementary approach to direct detection of the SARS-CoV-2 virus and holds potential for overcoming the limitations of existing COVID-19 diagnostics. RNA-sequencing (RNA-seq) studies of NP swabs and blood have demonstrated that COVID-19 elicits a unique host transcriptional response compared with non-viral and other viral acute respiratory illnesses (ARIs)^10–12^. A host gene expression signature of COVID-19, when utilized in combination with molecular detection of SARS-CoV-2, can serve as a fallback to identify suspected false negative or false positive results of traditional viral PCR tests, thus improving overall diagnostic reliability.

Recent studies have employed machine learning on RNA-seq data from NP swabs to develop proof-of-concept, host-based COVID-19 diagnostic classifiers that rely on a relatively large number of genes^10,13^. While highly promising, these classifiers have yet to undergo validation in external cohorts. Furthermore, RNA-seq is not widely available in clinical settings and thus the immediate practical utility of RNA-seq classifiers is limited.

Here, we address these gaps by developing 2-gene host signatures that could practically be incorporated into an RT-qPCR (qPCR) assay alongside a control gene and a viral target. We leverage NP swab RNA-seq data from two large patient cohorts to derive and validate top performing support vector machine (SVM) binary classifiers that use 2 host genes to predict COVID-19 status. We then refine these signatures for use in qPCR assays and confirm their prediction performance from qPCR data on an independent sample cohort.

The optimal 2-gene signatures combine an interferon-stimulated gene (ISG) that is strongly induced in COVID-19, such as *IFI6, IFI44* or *IFI44L*, with another immune response gene that is more strongly induced in other viral ARIs, such as *GBP5* or *CCL3*. Finally, we demonstrate that such a host classifier is robust across SARS-CoV-2 variants, including those that can yield a false negative viral PCR result, and is unaffected by laboratory cross-contamination that can yield a false positive viral PCR result.

## Results

### Performance of 2-gene combinations for a COVID-19 host classifier from NP swab RNA-seq

We previously developed multi-gene host classifiers for COVID-19 using RNA-seq data from NP swabs of patients tested for COVID-19 at the University of California, San Francisco (UCSF) who were diagnosed with either COVID-19, other viral ARIs or non-viral ARIs^10^. In the present work, we sought to develop a parsimonious 2-gene signature that could practically be incorporated into a PCR test alongside a control gene and a viral target. We began by identifying top performing 2-gene candidates in our RNA-seq cohort after supplementing it with additional samples collected in the intervening time. The full UCSF cohort included n=318 patients, of whom 90 had COVID-19 (with viral load equivalent to PCR C_t_ < 30), 59 had other viral ARIs (mostly rhinovirus and influenza), and 169 had non-viral conditions (**Supp. Table 1**; **Supp. Data File 1**).

The UCSF samples were split into a training set (70%) and a testing set (30%), with stratification to ensure each one contained similar proportions of samples with and without COVID-19. We then applied a greedy selection algorithm to identify 2-gene combinations that best predicted COVID-19 status. The performance metric was the area under the receiver operating characteristic curve (AUC) of a support vector machine (SVM) binary classifier that used the selected genes as features, calculated using 5-fold cross-validation within the training set (**Figure 1a**). Thus, a first gene was selected to maximize the AUC it achieved, and a second gene was selected to maximize the AUC when combined with the first gene. **Table 1a** lists nine combinations composed of each of the three best ‘first’ genes and their respective three best ‘second’ genes. The ‘first’ genes in the top combinations were the interferon-stimulated genes (ISGs) *IFI6, IFI44L* and *HERC6*, which we previously showed are strongly induced in COVID-19^10^. Most of the ‘second’ genes were also related to immune and inflammatory processes.

**Table 1.**
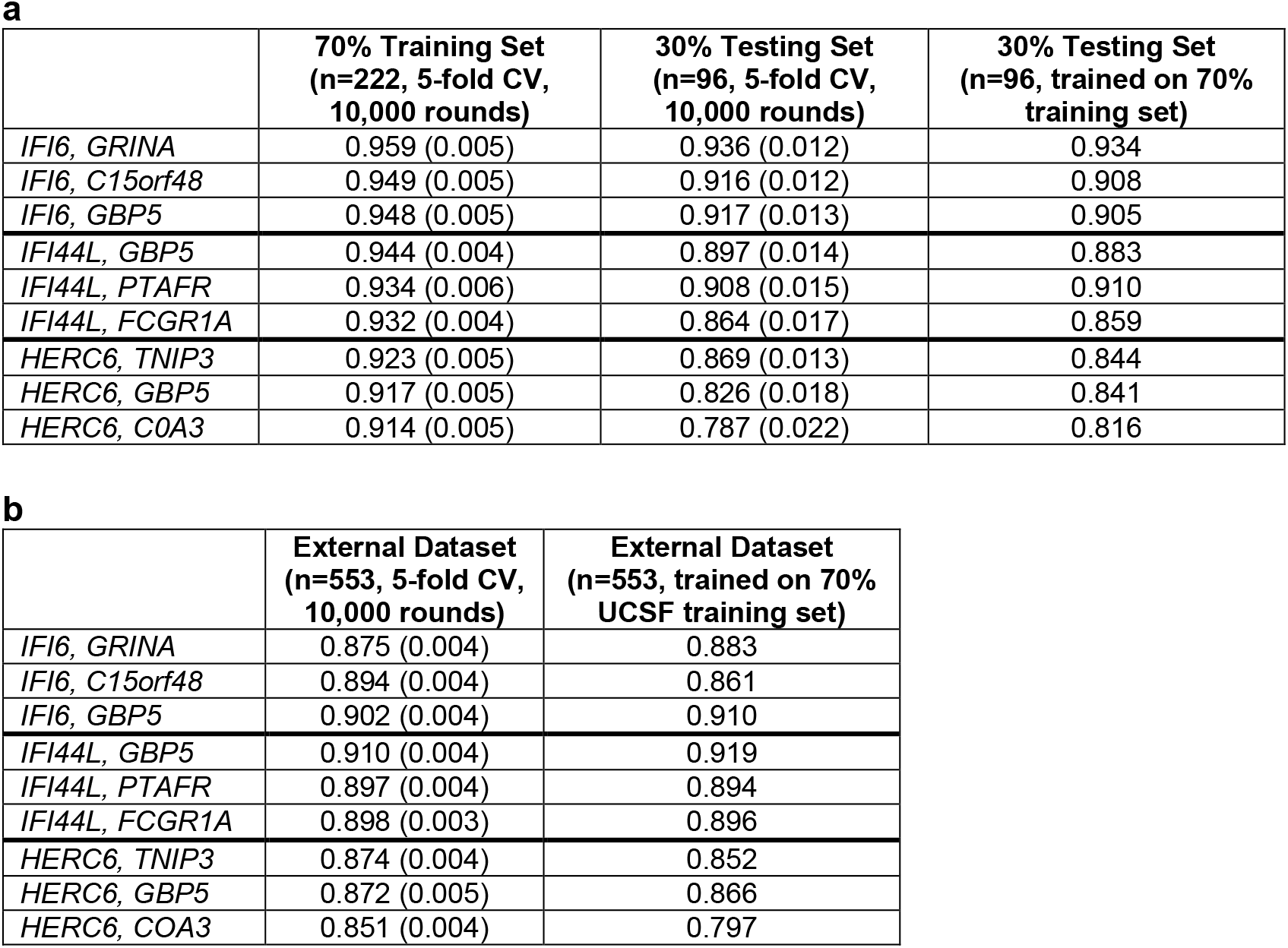
Performance of 2-gene SVM classifiers on RNA-seq data. Performance of binary SVM classifiers for predicting COVID-19 status, measured by the area under the curve (AUC). Where multiple cross-validation (CV) rounds were performed, values indicate mean and standard deviation. **a)** Performance of indicated 2-gene combinations in the UCSF training (70%) and testing (30%) sets, **b)** Performance in the external New York dataset.

|  | 70% Training Set<br>(n=222, 5-fold CV,<br>10,000 rounds) | 30% Testing Set<br>(n=96, 5-fold CV,<br>10,000 rounds) | 30% Testing Set<br>(n=96, trained on 70%<br>training set) |
| --- | --- | --- | --- |
| *IFI6, GRINA* | 0.959 (0.005) | 0.936 (0.012) | 0.934 |
| *IFI6, C15orf48* | 0.949 (0.005) | 0.916 (0.012) | 0.908 |
| *IFI6, GBP5* | 0.948 (0.005) | 0.917 (0.013) | 0.905 |
| *IFI44L, GBP5* | 0.944 (0.004) | 0.897 (0.014) | 0.883 |
| *IFI44L, PTAFR* | 0.934 (0.006) | 0.908 (0.015) | 0.910 |
| *IFI44L, FCGR1A* | 0.932 (0.004) | 0.864 (0.017) | 0.859 |
| *HERC6, TNIP3* | 0.923 (0.005) | 0.869 (0.013) | 0.844 |
| *HERC6, GBP5* | 0.917 (0.005) | 0.826 (0.018) | 0.841 |
| *HERC6, COA3* | 0.914 (0.005) | 0.787 (0.022) | 0.816 |

**b**
|  | External Dataset<br>(n=553, 5-fold CV,<br>10,000 rounds) | External Dataset<br>(n=553, trained on 70%<br>UCSF training set) |
| --- | --- | --- |
| *IFI6, GRINA* | 0.875 (0.004) | 0.883 |
| *IFI6, C15orf48* | 0.894 (0.004) | 0.861 |
| *IFI6, GBP5* | 0.902 (0.004) | 0.910 |
| *IFI44L, GBP5* | 0.910 (0.004) | 0.919 |
| *IFI44L, PTAFR* | 0.897 (0.004) | 0.894 |
| *IFI44L, FCGR1A* | 0.898 (0.003) | 0.896 |
| *HERC6, TNIP3* | 0.874 (0.004) | 0.852 |
| *HERC6, GBP5* | 0.872 (0.005) | 0.866 |
| *HERC6, COA3* | 0.851 (0.004) | 0.797 |

**Figure 1.**
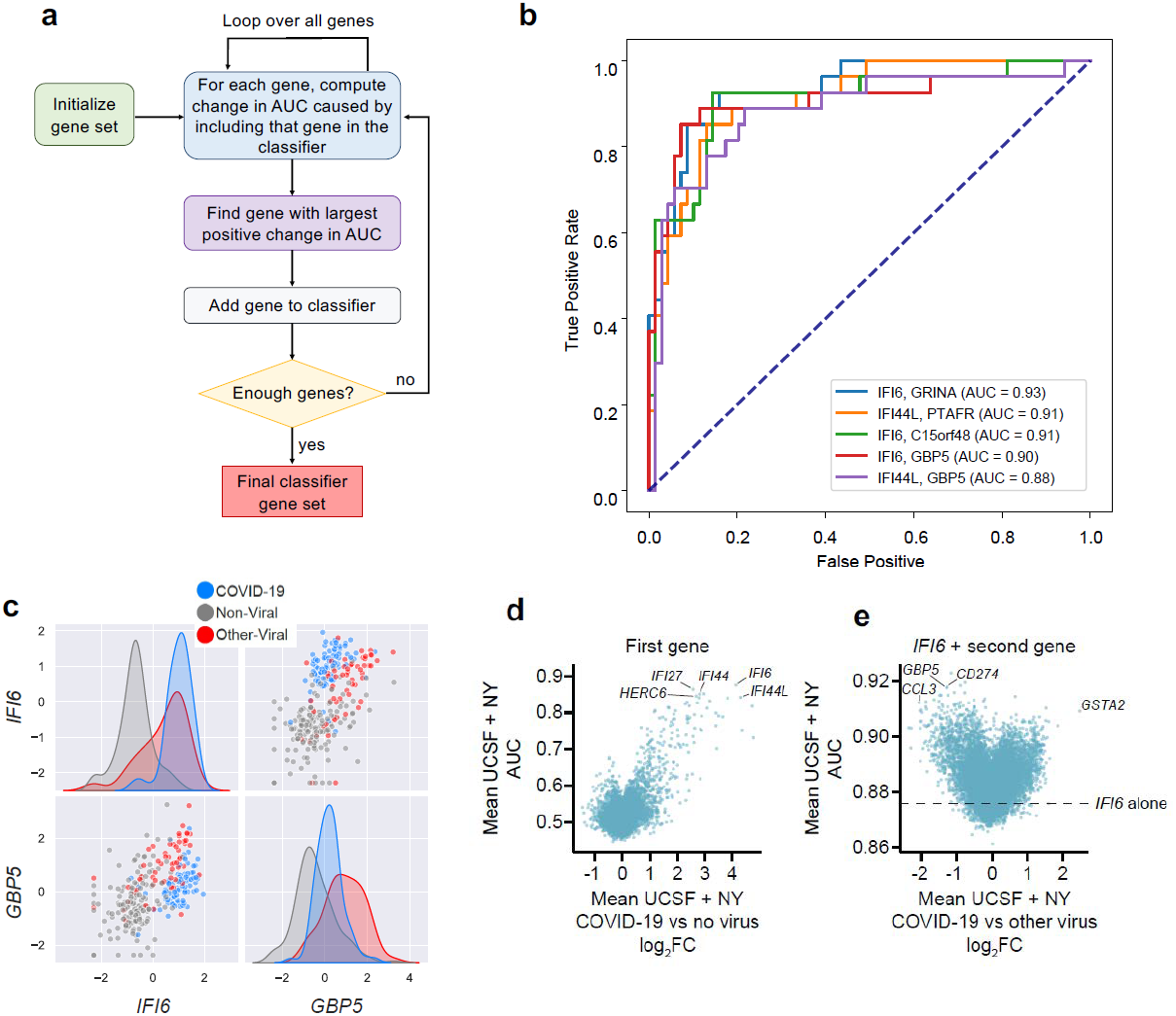
Development of 2-gene host-based SVM COVID-19 diagnostic classifiers from RNA-seq data. **a)** Schematic of the greedy feature selection algorithm used to identify top performing 2-gene combinations. **b)** Receiver operating characteristic (ROC) curve demonstrating performance of SVM classifiers using the indicated 2-gene combinations. The classifiers were trained on the UCSF training set and applied to the UCSF testing set. AUC = area under the ROC curve. **c)** Expression scatter plots and distributions of the representative ‘first’ and ‘second’ genes *IFI6* and *GBP5*, respectively, in the full UCSF cohort. Shown are variance-stabilized gene expression values following standardization. Color indicates patient group. **d)** Scatter plot of the AUC of COVID-19 diagnostic classifiers that rely on each single gene, calculated using 5-fold cross-validation (y-axis), against log_2_ fold-change (log_2_FC) of the gene between the COVID-19 and non-viral samples (x-axis). Both metrics were averaged between the full UCSF cohort and the New York cohort. **e)** Scatter plot of the AUC of COVID-19 diagnostic classifiers that rely on the combination of *IFI6* and each possible ‘second’ gene, calculated using 5-fold cross-validation (y-axis), against log2 fold-change (log2FC) of the ‘second’ gene between the COVID-19 and other viral samples (x-axis). Both metrics were averaged between the full UCSF cohort and the New York cohort. The AUC of an *IFI6*-only classifier is shown for reference.

The performance of the nine 2-gene combinations on previously unseen data was estimated by: i) 10,000 rounds of 5-fold cross-validation within the training set, ii) 10,000 rounds of 5-fold cross-validation within the testing set, or iii) training on the training set and prediction on the testing set (**Table 1a**). Using the third approach, we observed AUC values as high as 0.93 (**Figure 1b**). We further validated the classifiers using an external, independently generated and quantified NP swab RNA-seq dataset from a cohort of n=553 patients in New York (166 with COVID-19, 79 with other viral ARIs, 308 with non-viral conditions)^12^ (**Supp. Table 1**; **Supp. Data File 1**). The 2-gene combinations achieved comparable performance on the external dataset (**Table 1b**). The best performing combinations were *IFI44L*+*GBP5* (AUC 0.919) and *IFI6*+*GBP5* (AUC 0.91), when the classifier was trained on the UCSF 70% training set. These results demonstrate that 2-gene classifier models are feasible, stable, and generalizable.

### Optimization of 2-gene combinations for incorporation into an RT-qPCR assay

We noted that the ‘first’ and the ‘second’ genes in the 2-gene combinations performed distinct roles. The former was sufficient to distinguish COVID-19 from non-viral ARIs while the latter helped reinforce the distinction between COVID-19 and other viral ARIs. Considering *IFI6* and *GBP5* as an example, *IFI6* alone almost completely separated the COVID-19 and non-viral samples (**Figure 1c**). However, some of the other viral ARI samples showed equivalent levels of *IFI6* expression. Adding *GBP5* to the model allowed for improved separation, as expression of this ISG was typically higher in other viral ARIs (**Figure 1c**). Given this pattern, and because our ultimate goal was a qPCR assay in which small effect sizes are more difficult to discern, we refined our candidate genes by also considering the expression fold-change between COVID-19 and the two other patient groups.

We first plotted the AUC of SVM classifiers relying on each individual gene against the fold-change of that gene between the COVID-19 and non-viral samples, where both measures were averaged between the full UCSF cohort and the New York cohort (**Figure 1d**; **Supp. Data File 2**). As expected, several ISGs exhibited equivalently robust predictive value as well as substantial fold-changes (log_2_FC ∼2-4) that should be readily detectable by qPCR. We then plotted the AUC of classifiers that used the ISG *IFI6* in combination with every possible ‘second’ gene, against the fold-change of the ‘second’ gene between COVID-19 and other viral ARIs (**Figure 1e**; **Supp. Data File 2**). This revealed candidate genes with somewhat smaller fold-changes (log_2_FC ∼1.5-2) that should still be detectable by qPCR. These candidate genes only partly overlapped with the ‘second’ genes selected by the greedy algorithm, which did not explicitly consider fold-change.

### RT-qPCR validation of host genes to differentiate COVID-19 from other ARIs

We chose four ‘first’ ISGs based on their predictive value and fold-change. We then measured the expression of these ISGs, relative to the reference gene *RPP30*, using qPCR in swabs from a new cohort of patients with (n=72) or without (n=72) COVID-19. Because these swabs were not sequenced, we could not definitively assign those without COVID-19 as either non-viral or other viral cases. However, the low prevalence of other viral ARIs during the timeframe of sample collection, due to the public health measures implemented for COVID-19^14^, suggested they were mostly non-viral. All four genes were able to clearly separate the majority of samples with or without COVID-19 in the qPCR data (**Figure 2a**). SVM classifiers relying on single ISGs achieved mean AUC values as high as 0.88 in predicting COVID-19 status from the qPCR data (**Figure 2b**), on par with their prediction performance from RNA-seq (**Figure 1d**).

**Figure 2.**
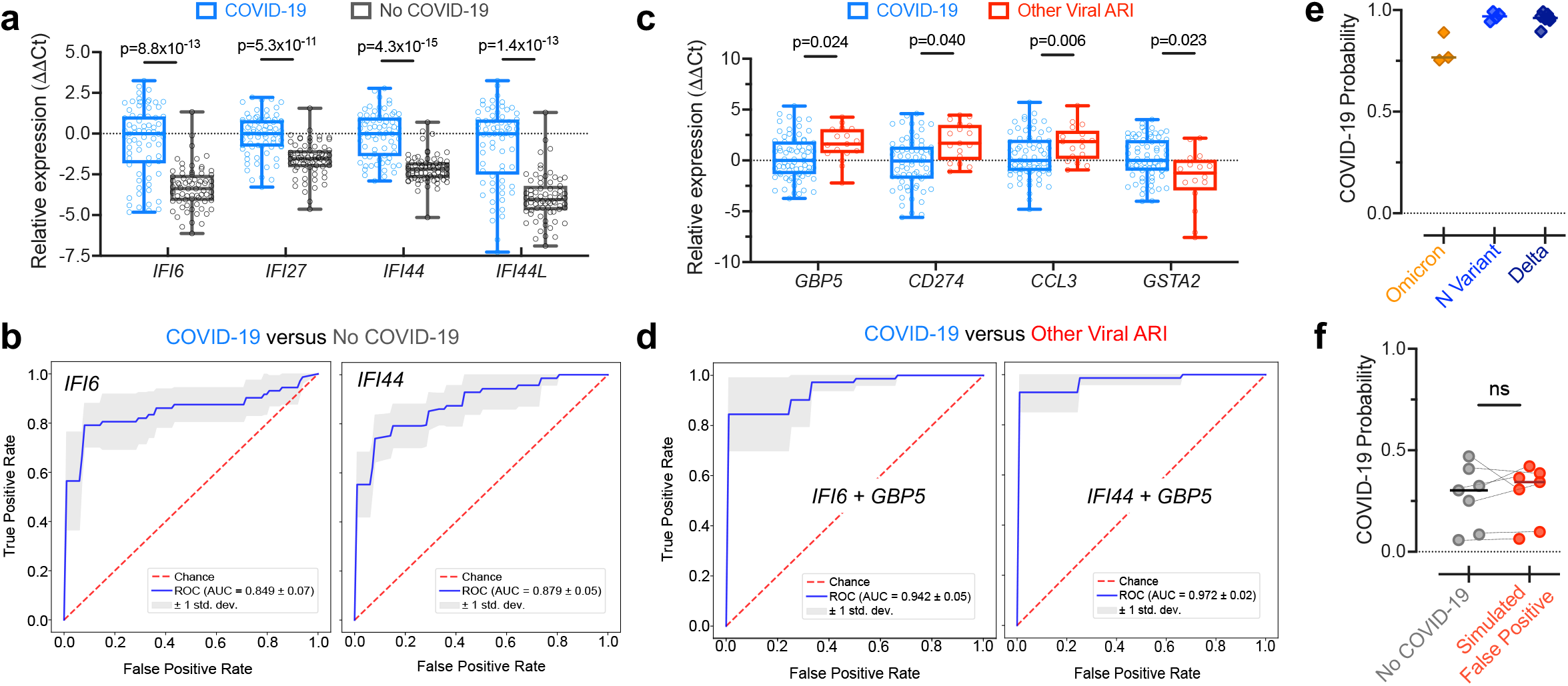
Performance of 2-gene SVM COVID-19 diagnostic classifiers in qPCR assays. **a)** Expression differences determined by qPCR in a new cohort of patients with (n=72) or without (n=72) COVID-19 for several ‘first’ ISGs selected for their predictive value and fold-change in the RNA-seq data. Shown are boxplots of ΔCt values, using *RPP30* as the reference gene, normalized to the median of the COVID-19 group. Statistical significance was assessed using a one-sided Mann-Whitney test with Bonferroni correction. **b)** ROC curves demonstrating performance of SVM classifiers relying on single ISGs for distinguishing the samples with or without COVID-19 using the qPCR data, estimated by 5-fold cross-validation. **c)** Expression differences determined by qPCR between the samples with COVID-19 (n=72) and samples with other viral ARIs (n=17; a subset of the samples from Figure 1) for several ‘second’ genes selected for their predictive value and fold-change in the RNA-seq data. Shown are boxplots of ΔCt values, using *RPP30* as the reference gene, and normalized to the median of the COVID-19 group. Statistical significance was assessed using a one-sided Mann-Whitney test with Bonferroni correction. **d)** ROC curves demonstrating performance of SVM classifiers relying on 2-gene combinations for distinguishing samples with COVID-19 from samples with other viral ARIs using the qPCR data, estimated by 5-fold cross-validation. **e)** Probability of COVID-19 predicted by the *IFI6*+*GBP5* classifier using the qPCR data for samples with the Omicron variant (n=3), the N-gene variant (n=4), and the Delta variant (n=7). The classifier was trained on the samples with and without COVID-19 shown in a). **f)** Probability of COVID-19 predicted by the *IFI6*+*GBP5* classifier using the qPCR data for n=7 samples without COVID-19 before and after trace contamination from a sample with high SARS-CoV-2 viral load. The classifier was trained on the samples with and without COVID-19 shown in a). Statistical significance was assessed using a one-sided, paired Mann-Whitney test. ns = P > 0.05, * = P < 0.05, ** = P < 0.01, *** = P < 0.001, **** = P < 0.0001.

To explicitly test the ability to separate COVID-19 from other respiratory viruses using qPCR data, we chose four ‘second’ genes and measured their expression in the COVID-19 samples described above (n=72) as compared to a subset of our original, sequenced other viral samples (n=17). Consistent with the RNA-seq data, the expression of three of the genes (*GBP5, CD274* and *CCL3*) was significantly higher on average in the other viral samples, and expression of the fourth gene (*GSTA2*) was significantly higher in the COVID-19 samples (**Figure 2c**). Classifiers that combined one of these genes with a ‘first’ gene achieved near perfect separation of the COVID-19 and other viral samples (**Figure 2d**). This performance is likely overly optimistic, due in part to the relatively small size of the other virus group in the qPCR data, but it is overall consistent with the performance observed in the larger RNA-seq datasets. These results demonstrate that 2-gene signatures can successfully predict COVID-19 status from qPCR data.

### Host signatures are robust to SARS-CoV-2 variants and laboratory cross-contamination

We next assessed whether a 2-gene host classifier was robust across SARS-CoV-2 variants, which could conceivably yield an altered host response and/or harbor mutations that disrupt primer target sites and lead to false negative viral PCR tests^5,7,15^. We performed qPCR for the genes *IFI6* and *GBP5* on samples with the Omicron variant (n=3), which causes S-gene target dropout in certain viral PCR assays; the N-gene variant (n=4), which causes N-gene target dropout^15^; and on samples with the Delta variant (n=7). An SVM classifier trained on the qPCR results of the samples with and without COVID-19, described above, predicted COVID-19 with high likelihood in all variant samples (**Figure 2e**), demonstrating the utility of a host signature as a complement to viral PCR.

On the other hand, false positive viral PCR tests frequently result from trace cross-contamination of samples with high viral titers into negative specimens processed contemporaneously in the laboratory^9^. To examine whether the *IFI6*+*GBP5* host classifier would be affected in such cross-contamination events, we spiked extracted NP swab RNA from a sample with very high SARS-CoV-2 viral load (C_t_≈12) into n=7 COVID-19 negative swab specimens at a dilution of 1:10^5^, which would be expected to yield a positive viral PCR with C_t_<30. The probability of COVID-19 estimated by the host classifier was not significantly affected in the simulated false-positive specimens (**Figure 2f**).

## Discussion

We leveraged multiple cohorts – encompassing over 1,000 patients with COVID-19, other viral ARIs and non-viral conditions – to develop and validate 2-gene host-based COVID-19 diagnostic classifiers that could be practically incorporated into clinical PCR assays in combination with a control gene and a viral target. We found that the host classifier enabled reliable identification of COVID-19 even in the face of SARS-CoV-2 variants that cause false negative viral PCR tests and remained unaffected by simulated laboratory cross-contamination that can cause false positive viral PCR tests.

Given the inevitable continued emergence of SARS-CoV-2 variants, which may disrupt primer target sites, assays capable of detecting infection regardless of viral sequence are essential to avoid adverse outcomes owing to infected individuals going unrecognized in congregate settings, such as hospitals or nursing homes. The adverse effects of false-positive tests are also non-trivial. The positive predictive value of highly specific viral PCR assays diminishes for asymptomatic individuals undergoing continual surveillance testing in low prevalence settings^9^. False positive results then become more likely, leading to unnecessary isolation and quarantine, depletion of essential personnel, and unwarranted contact tracing.

Our study has some limitations. While our findings provide a framework for the rapid clinical translation of a host-based COVID-19 diagnostic, a randomized controlled trial of our assay will be needed to firmly establish its clinical utility. Our results suggest that addition of host targets is likely to improve diagnostic accuracy, however, a prospective assessment using clinically confirmed false-positive and false-negative viral tests is needed. Moreover, our classifier models were trained and tested on cohorts with particular characteristics, including the balance between COVID-19, other viral and non-viral samples; the mix of other respiratory viruses represented; and within the COVID-19 group, the distributions of viral load and of time since onset of infection. All these variables no doubt affect classifier performance and will vary in reality with time and place. However, the fact that our classifiers translated so well across diverse real-world cohorts argues that they are quite robust to these issues.

While we did not explicitly explore it here, our results suggest that parsimonious host classifiers could serve not only as a COVID-19 diagnostic but also as a pan-respiratory virus surveillance tool. Even prior to the COVID-19 pandemic, viral lower respiratory tract infections were a leading cause of disease and death^16^, and many respiratory viral infections go undetected, leading to preventable transmission and unnecessary antibiotic treatment^17^. Since our classifiers rely heavily on ISGs, and type I interferon signaling is a biologically conserved mechanism, these genes could be used in future work as the basis for a diagnostic that identifies respiratory viruses more generally. Such a diagnostic could have considerable value as a screening tool in hospitals, nursing homes or other congregate settings with potential for adverse consequences from unrecognized respiratory viral transmission.

## Materials and Methods

### RNA-seq cohorts and data pre-processing

The UCSF cohort used to develop the RNA-seq classifiers was initially described in our study applying metagenomic sequencing to NP swabs from adult patients tested for COVID-19 by RT-PCR, according to the published methods and under UCSF IRB #17-24056^10^. In brief, samples were assigned to one of three viral status groups: 1) samples with a positive clinical RT-PCR test for SARS-CoV-2 were assigned to the “COVID-19” group, 2) samples with another pathogenic respiratory virus detected by the ID-Seq pipeline^18^ in the metagenomic sequencing data were assigned to the “other virus” group, and 3) remaining samples were assigned to the “no virus” group. In the present work, we supplemented the samples reported in our original study with additional swabs collected, sequenced and analyzed in the same manner.

We wished to retain for classifier development COVID-19 samples with likely active infection (culturable virus), which several studies have related to viral PCR C_t_ < 30^19–21^. Because not all C_t_ values were available, we relied on the relationship between viral reads-per-million (rpM) in the sequencing data and PCR C_t_ that we previously reported^10^: log_2_(rpM) = 31.9753 - 0.9167*C_t_. Metadata for the UCSF samples is provided in **Supp. Data File 1**.

We pseudo-aligned the UCSF samples with kallisto^22^ (v. 0.46.1), using the bias correction setting, against an index consisting of all transcripts associated with human protein coding genes (ENSEMBL v. 99), cytosolic and mitochondrial ribosomal RNA sequences, and the sequences of ERCC RNA standards. Samples retained in the dataset had at least 400,000 estimated counts associated with transcripts of protein coding genes. Gene-level counts were generated from the kallisto transcript abundance estimates using the R package tximport^23^ (v. 1.14) with the scaledTPM method. Genes were retained if they had at least 10 counts in at least 20% of samples.

The New York cohort used to validate the RNA-seq SVM classifiers on an external dataset was previously published^12^. Samples in this cohort were also categorized into the three viral status groups described above based on a combination of RT-PCR and metagenomic sequencing. Because we did not have access to the underlying sequencing data, we used the gene counts originally generated by the authors using STAR alignment and the R function featureCounts. We excluded samples with less than five million total counts as well as samples that had discordant COVID-19 test results between two assays, but did not filter based on viral load. Genes were retained if they had at least 32 counts in at least 10% of samples. Metadata for the New York samples is provided in **Supp. Data File 1**.

For each RNA-seq cohort, gene counts were subjected to the variance stabilizing transformation (VST) from the R package DESeq2 (v. 1.26.0) and the transformed values were then standardized (centered and scaled) to yield the final input features.

### RNA-seq SVM classifier development and validation

SVM learning was implemented in scikit-learn (https://scikit-learn.org) using the sklearn.svm.SVC class function with default parameters and probabilistic output.

The UCSF cohort was split into a training set (70%) and a testing set (30%), with stratification to ensure each set contained a similar proportion of samples with and without COVID-19. For the greedy feature selection, performance of a binary SVM classifier for predicting COVID-19 status relying on each single feature (gene) was evaluated by running 5-fold cross-validation within the training set and calculating the average AUC across the folds. The three best-performing ‘first’ genes were then selected. To extend these ‘first’ genes to 2-gene combinations, another round of the algorithm was performed, picking the three best-performing ‘second’ genes when combined with each of the ‘first’ genes.

In order to rigorously assess the performance of the SVM 2-gene models, we employed three approaches: (1) running 10,000 rounds of 5-fold cross-validation on the UCSF 70% training set and calculating the average AUC and standard deviation, (2) running 10,000 rounds of 5-fold cross-validation on the UCSF 30% testing set and calculating the average AUC and standard deviation, and (3) training each model on the UCSF 70% training set and testing it on the 30% testing set to generate an AUC score (**Table 1a**). We then validated the 2-gene models on the external New York cohort, using two approaches: (1) running 10,000 rounds of 5-fold cross-validation on the New York cohort and calculating the average AUC and standard deviation, and (2) training each model on the UCSF 70% training set and testing it on the New York cohort to generate an AUC score (**Table 1b**).

The AUC scores of single gene or 2-gene SVM classifiers displayed in **Figure 1d,e** were calculated by 5-fold cross validation using all the samples in each RNA-seq cohort.

### RNA-seq differential expression

Gene expression fold-changes in each RNA-seq cohort between the COVID-19 and non-viral samples (**Figure 1d**) and between the COVID-19 and other viral samples (**Figure 1e**) were calculated with the R package limma (v. 3.42), using quantile normalization and the voom method.

### RT-qPCR of host genes

RNA was reverse transcribed using the High-Capacity cDNA Reverse Transcription Kit (Applied Biosystems), according to the manufacturer’s protocol, and analyzed by qPCR in a Bio-Rad CFX384 thermocycler (BioRad) using Taqman Fast Advanced Master Mix (Applied Biosystems) and Taqman Gene Expression Assays (Applied Biosystems), according to the manufacturer’s protocol. Assay IDs for each gene are provided in **Supp. Table 2**. ΔC_t_ values were calculated with respect to the reference gene *RPP30* (also known as *RNASEP2*), the standard host control gene used in many viral PCR tests. ΔC_t_ values are provided in **Supp. Data File 3**.

### qPCR SVM classifier development and validation

The input features for qPCR-based SVM COVID-19 diagnostic classifiers were standardized (centered and scaled) ΔC_t_ values. Standardization was performed separately in each analyzed sample set using the mean and standard deviation of the training samples. In the context of cross-validation, this was done for each fold using the appropriate training samples. Performance of SVM classifiers to distinguish between the samples with (n=72) and without (n=72) COVID-19 (**Figure 2b**), or to distinguish between the samples with COVID-19 and other viral ARIs (n=17) (**Figure 2d**), was assessed by 5-fold cross-validation.

The *IFI6*+*GBP5* classifier, which was used to predict the COVID-19 status of variant samples (**Figure 2e**) and of samples that had been purposely contaminated with 1:10^5^ dilution from a high SARS-CoV-2 viral load sample (**Figure 2f**), was trained on the set of samples with and without COVID-19 described above. Because the variant and contamination samples on which we performed prediction were assayed in separate experiments subsequent to the generation of the training dataset, they were always processed alongside n=6-7 COVID-19 negative controls from the original training dataset. The median ΔC_t_ difference observed for these control samples between the training dataset and the prediction experiment in which they were re-run was applied to all the samples in the respective experiment in order to account for systematic shifts.

## Data Availability

Gene counts for all UCSF samples have been deposited under NCBI GEO accession GSE188678. The New York dataset can be obtained according to the Data Availability statement in the original publication12. Code for RNA-seq and qPCR SVM classifier development and validation is available at: https://github.com/czbiohub/Covid-Host-Classifier-Code.

## Data Availability

Gene counts for all UCSF samples have been deposited under NCBI GEO accession GSE188678. The New York dataset can be obtained according to the Data Availability statement in the original publication^12^. Code for RNA-seq and qPCR SVM classifier development and validation is available at: https://github.com/czbiohub/Covid-Host-Classifier-Code.

## Supplementary Tables

**Supplementary Table 1.** Cohort details.

| Cohort | Description | COVID-19 (n) | Non-Viral ARI (n) | Other Viral ARI (n) | Reference |
| --- | --- | --- | --- | --- | --- |
| 1. UCSF (RNA-seq, n=318) | Patients tested for COVID-19, CA, 2020 | 90 | 169 | 59 | <sup>10</sup> + this study |
| 2. New York (RNA-seq, n=553) | Patients tested for COVID-19, NY, 2020 | 166 | 308 | 79 | <sup>12</sup> |
| 3. UCSF (qPCR, n=144) | Patients tested for COVID-19, CA, 2020 | 72 | 72 |  | This study |
| 4. UCSF SARS-CoV-2 N-gene variant (qPCR, n=4) | Patients with SARS-CoV-2 N-gene variant, CA, 2020 | 4 | - |  | This study |
| 5. UCSF SARS-CoV-2 Delta variant (qPCR, n=7) | Patients with SARS-CoV-2 Delta variant, CA, 2021 | 7 | - |  | This study |
| 6. UCSF SARS-CoV-2 Omicron variant (qPCR, n=3) | Patients with SARS-CoV-2 Omicron variant, CA, 2021 | 3 | - |  | This study |

**Supplementary Table 2.** Taqman Gene Expression assay IDs for genes tested by RT-qPCR.

| Gene | Assay ID |
| --- | --- |
| *IFI6* | Hs00242571_m1 |
| *GBP5* | Hs00369472_m1 |
| *RPP30* | Hs01124518_m1 |
| *IFI44* | Hs00197427_m1 |
| *IFI44L* | Hs00915292_m1 |
| *IFI27* | Hs01086373_g1 |
| *CCL3* | Hs00234142_m1 |
| *CD274* | Hs00204257_m1 |
| *GSTA2* | Hs00747232_mH |

## Supplementary Data Files

**Supplementary Data File 1**. Sample metadata for the UCSF and New York RNA-seq cohorts.

**Supplementary Data File 2**. AUC values and expression fold-changes in the UCSF cohort and in the New York cohort for every possible ‘first’ gene and for every possible ‘second’ gene combined with *IFI6* (related to **Figure 1d,e**).

**Supplementary Data File 3**. ΔC_t_ values for host genes in all the samples used in qPCR assays (related to **Figure 2a,c,e,f**). Clinical RT-PCR SARS-CoV-2 C_t_ values are also provided for the COVID-19 positive samples.

## References

1. World Health Organization. WHO COVID-19 Dashboard. https://covid19.who.int (2021).

2. Arevalo-Rodriguez, I. et al. False-negative results of initial RT-PCR assays for COVID-19: A systematic review. PLoS ONE 15, e0242958 (2020).

3. Long, D. R. et al. Occurrence and Timing of Subsequent Severe Acute Respiratory Syndrome Coronavirus 2 Reverse-transcription Polymerase Chain Reaction Positivity Among Initially Negative Patients. Clinical Infectious Diseases 72, 323–326 (2021).

4. Kanji, J. N. et al. False negative rate of COVID-19 PCR testing: a discordant testing analysis. Virol J 18, 13 (2021).

5. van Dorp, L. et al. Emergence of genomic diversity and recurrent mutations in SARS-CoV-2. Infection, Genetics and Evolution 83, 104351 (2020).

6. Galloway, S. E. et al. Emergence of SARS-CoV-2 B.1.1.7 Lineage — United States, December 29, 2020–January 12, 2021. MMWR Morb. Mortal. Wkly. Rep. 70, 95–99 (2021).

7. U.S. Food and Drug Administration. Genetic Variants of SARS-CoV-2 May Lead to False Negative Results with Molecular Tests for Detection of SARS-CoV-2 -Letter to Clinical Laboratory Staff and Health Care Providers. https://www.fda.gov/medical-devices/letters-health-care-providers/genetic-variants-sars-cov-2-may-lead-false-negative-results-molecular-tests-detection-sars-cov-2 (2021).

8. Surkova, E., Nikolayevskyy, V. & Drobniewski, F. False-positive COVID-19 results: hidden problems and costs. The Lancet Respiratory Medicine 8, 1167–1168 (2020).

9. Healy, B., Khan, A., Metezai, H., Blyth, I. & Asad, H. The impact of false positive COVID-19 results in an area of low prevalence. Clin Med 21, e54–e56 (2021).

10. Mick, E. et al. Upper airway gene expression reveals suppressed immune responses to SARS-CoV-2 compared with other respiratory viruses. Nature Communications 11, (2020).

11. McClain, M. T. et al. Dysregulated transcriptional responses to SARS-CoV-2 in the periphery. Nat Commun 12, 1079 (2021).

12. Butler, D. et al. Shotgun transcriptome, spatial omics, and isothermal profiling of SARS-CoV-2 infection reveals unique host responses, viral diversification, and drug interactions. Nat Commun 12, 1660 (2021).

13. Ng, D. L. et al. A diagnostic host response biosignature for COVID-19 from RNA profiling of nasal swabs and blood. Sci. Adv. 7, eabe5984 (2021).

14. Olsen, S. J. et al. Changes in Influenza and Other Respiratory Virus Activity During the COVID-19 Pandemic — United States, 2020–2021. MMWR Morb. Mortal. Wkly. Rep. 70, 1013–1019 (2021).

15. Vanaerschot, M. et al. Identification of a Polymorphism in the N Gene of SARS-CoV-2 That Adversely Impacts Detection by Reverse Transcription-PCR. J Clin Microbiol 59, (2020).

16. World Health Organization. The top 10 causes of death. (2020).

17. Chow, E. J. & Mermel, L. A. Hospital-Acquired Respiratory Viral Infections: Incidence, Morbidity, and Mortality in Pediatric and Adult Patients. Open Forum Infect Dis 4, ofx006 (2017).

18. Kalantar, K. L. et al. IDseq—An open source cloud-based pipeline and analysis service for metagenomic pathogen detection and monitoring. Gigascience 9, (2020).

19. Singanayagam, A. et al. Duration of infectiousness and correlation with RT-PCR cycle threshold values in cases of COVID-19, England, January to May 2020. Eurosurveillance 25, (2020).

20. Bullard, J. et al. Predicting Infectious Severe Acute Respiratory Syndrome Coronavirus 2 From Diagnostic Samples. Clinical Infectious Diseases 71, 2663–2666 (2020).

21. La Scola, B. et al. Viral RNA load as determined by cell culture as a management tool for discharge of SARS-CoV-2 patients from infectious disease wards. Eur J Clin Microbiol Infect Dis 39, 1059–1061 (2020).

22. Bray, N. L., Pimentel, H., Melsted, P. & Pachter, L. Near-optimal probabilistic RNA-seq quantification. Nature Biotechnology 34, 525 (2016).

23. Soneson, C., Love, M. I. & Robinson, M. D. Differential analyses for RNA-seq: transcript-level estimates improve gene-level inferences. F1000Research 4, 1521 (2015).

